# Evolving clinical roles in the era of genomic medicine: Insights from rapid genome sequencing in the neonatal intensive care unit

**DOI:** 10.64898/2026.09.28.26363665

**Authors:** Peter Taber, Chelsea Solorzano, L Weaver, Kelsey A. Simek, Rachel Palmquist, Jorie Butler, Tanner Ellsworth, Mickey Bolyard, Martin Tristani-Firouzi, Paul Estabrooks, Sabrina Malone Jenkins

## Abstract

**Background:** Rapid genome sequencing (rGS) is a first-tier test in the neonatal intensive care unit (NICU). Implementation strategies to scale rGS for clinical care may change clinician roles. We analyzed the NICU rGS workflow to understand challenges rGS poses for clinical roles and responsibilities.

**Methods:** The study took a qualitative approach to understanding the rGS workflow in Level III and Level IV NICUs. The five workflow phases studied included: i) identification of a patient with a suspected genetic disorder, ii) subspecialist consultation, iii) test selection and ordering, iv) return of results, and v) disclosure. Data were collected via job task journals, interviews, and workflow diagram construction with clinicians involved in rGS. Analysis included participant-guided workflow diagram refinement, qualitative thematic analysis, and identification of rGS workflow challenges.

**Results:** We documented issues related to decision-making authority and scope of responsibility for clinicians, with the majority relating to complexities of subspecialist consultations. Issues included differences in: criteria for triggering consultations, expectations of consulted subspecialists’ roles, test selection preferences, and responsibility for documenting clinical interpretations and disclosure.

**Conclusion:** Subspecialist consultations in clinical rGS involved significant role ambiguity and communication challenges, requiring intentional implementation efforts to ensure clear clinical roles and responsibilities.

## Background

As rapid genome sequencing (rGS) becomes standard of care in the neonatal intensive care unit (NICU)^1–3^, test volumes may strain existing care delivery models. Prior work on genomic medicine implementation highlights diverse implementation strategies to support effective and scalable implementation^4–9^ such as multi-disciplinary involvement in genetic testing workflows^10,11^, embedded clinical genetic counselors^12–14^, and training for advanced practice providers (APPs) and non-genetics subspecialists to streamline clinician involvement in genetic testing.^15–18^ These strategies may improve the reach, adoption, and feasibility of rGS by distributing tasks across a broader clinical team, including non-specialists.^4^ At the same time, these care delivery adaptations may also complicate clinical roles and responsibilities in the genetic testing workflow by changing who is expected to recognize candidates for testing, initiate consultations, order or interpret tests, communicate findings, and arrange follow-up.^11,13,19,20^

How implementation strategies reshape clinical roles and responsibilities once rGS is embedded in routine care remains understudied. Ambiguities related to clinicians’ responsibilities in areas like communication and documentation may affect not only workflow efficiency, but also key implementation outcomes like reach, adoption, implementation fidelity, and sustainability of rGS programs. To contribute toward addressing this gap, we analyze current workflows for rGS at Level III (non-tertiary, rural lower resourced) and Level IV (tertiary, metropolitan, higher resourced) NICUs in two major Utah healthcare systems,^8,9^ reporting qualitative findings related to ambiguities and tensions in how different clinical roles understand their responsibilities.^21^

## Materials and methods

### Overview

This study characterized challenges related to clinical roles in rGS using an iterative, multi-modal qualitative study design guided by dissemination and implementation science. The work emerges from two parent studies seeking to integrate novel genomic diagnostic tools and support the reach, adoption, implementation, and maintenance of rGS across two levels of NICUs: Level III and Level IV (the highest level, with 24-hour access to specialists). Parent projects drew on the integrated Promoting Action on Research in Health Services (i-PARIHS) framework^22^ to examine characteristics of the innovation, organizational context, clinical teams, and facilitation processes; and on Reach, Effectiveness, Adoption, Implementation and Maintenance (RE-AIM) framework^23^ to operationalize outcomes. In accordance with the Common Rule, the workflow analysis was reviewed by the University of Utah Institutional Review Board and deemed exempt (IRB_00165986).

Major phases of the rGS workflow studied here are as follows:

*i) Identification of a patient with a suspected genetic disorder:* Nurses, advanced practice providers (APPs) or neonatologists observe something unusual about a neonate, leading to suspicion of an underlying genetic disorder. The responsible attending then initiates the rGS workflow.
*ii) Initial subspecialist consultations:* An attending may seek guidance via informal “curbside” consultations to the pediatric subspecialist/medical genetics team. A formal consultation is requested by the attending, either to the genetics service or non-genetics subspecialists. The choice of genetics or non-genetics subspecialty is often determined by the perception of whether the patient’s phenotype relates primarily to a single organ system. Consultations for perceived “single system” or “pure” phenotypes are placed to the relevant non-genetics subspecialty (e.g. pediatric neurology or gastroenterology). Non-genetics subspecialists may request consultation from medical genetics, depending on their evaluation. NICU providers at Level III facilities must arrange telegenetics appointments to evaluate the patient if a genetics consulation is desired.
*iii) Test selection and ordering:* After the consulted subspecialist(s) evaluates the patient, a recommendation is made to the NICU regarding genetic test selection. The NICU retains formal authority over test selection but may defer the decision to the consulted subspecialist. A genetic counselor often assists with determination, consent and ordering.
*iv) Return of results from reference laboratory and clinical interpretation:* Preliminary positive results are communicated via phone from the reference laboratory to the on-service attending. The final laboratory report is emailed to genetic counselors. The report is forwarded to the patient’s care team by the genetic counselor. Unexpectedly complex results can trigger additional consultations and/or tests. The care team discusses and interprets the results and documents the clinical interpretation.
*v) Disclosure of results to family and arrangement for follow-up care:* Disclosure of results varies depending on presence of positive variants and/or variants of uncertain significant (VUS), secondary findings, patient disposition, patient complexity, and other factors. Inpatient disclosures can include a meeting with multiple members of the care team or sequential meetings with clinicians to provide specialty-specific information and counseling. Level III return of results may require another telegenetics appointment, if a genetics consultation was placed for the patient.

### Setting

This study was conducted within Intermountain Health (IH) and University of Utah Health (UHealth), two large healthcare systems in Utah which contain one IH Level IV NICU, 5 IH Level III NICUs, and one UHealth Level III NICU. The IH Level IV rGS pathway was initiated in 2019 and has sent approximately 310 inpatient rGS to date. IH Level III rGS was introduced in 2022, with those sites sending approximately 85 rGS to date. Clinical implementation at the UHealth Level III site began in 2023^24^, with approximately 70 rGS sent to date.

### Participants

The study was performed with neonatologists, APPS, pediatric subspecialists, genetic counselors, medical geneticists and nurses working in Level III and Level IV NICUs. Molecular geneticists from a commercial laboratory were included in the job task journal component of data collection (see below). Participants were purposively recruited by email based on their prior experience with rGS and work within the NICUs of interest, with the study striving to capture the perspectives of diverse clinical roles. Participants were recruited from NICUs in which some of the research team also worked, and were provided information about local rGS implementation efforts as part of the interview.

### Data collection

Data collection utilized job task journals and two rounds of workflow elicitation interviews. Job task journals collected via Qualtrics (Qualtrics, LLC, Provo, Utah) or REDCap^25^ provided a rapid overview of tasks and roles in rGS, informing development of a preliminary workflow diagram and interview guide.

Two rounds of interviews were conducted with one to three participants from the same clinical role per interview. Interview guides were constructed with input from clinical experts on the study team (Auth2, Auth4, Auth5, Auth7, Auth11). Interviews were conducted virtually by a PhD anthropologist (Auth1) and lasted roughly an hour. Interviews included elicitation of clinical responsibilities and genomics background, overview of each role’s rGS workflow, description of memorable rGS cases via critical incident technique, and evaluation of an electronic health record (EHR) embedded phenotyping tool (to be reported separately).

Workflow elicitation utilized a diagram constructed in Lucidchart (Lucid Software, South Jordan, Utah) as a prompt to facilitate feedback on interviewees’ current practices, challenges faced, and desired rGS workflow improvements. This process allowed participants to identify where existing clinical routines, local communication norms, staffing models, or differences between Level III and Level IV settings shaped how rGS tasks were assigned or coordinated. No formal modeling system was used because the project’s goal was to use the workflow diagram as a communication tool with clinicians. Iterative interviewing allowed the research team to “member check” inferences and interpretations with clinician participants as an integrated part of data collection and analysis.^26^ Finally, to characterize adaptations in the local rGS program, the team examined existing literature on clinical genetics care delivery models^4,7,27,28^, and consulted with neonatologist project advisors working in the study NICUs.

### Data analysi

Interviews were digitally recorded and professionally transcribed. Both rounds of interviews were included in thematic analysis. Coding took a hybrid deductive-inductive approach. Initial codes were derived from i-PARIHS constructs relevant to the parent project, including characteristics of the innovation (rGS), clinical and organizational context, innovation recipients, and facilitation processes. Inductive codes captured emergent topics, including professional authority, boundaries between clinical roles, differences of opinion, communication practices, and responsibility for documentation or disclosure. The full corpus was consensus-coded by members of the qualitative study team (Auth1, Auth2, Auth3, Auth8). Subsequent analysis by the lead coder (Auth1) examined role-related codes by rGS task and workflow phase. RE-AIM was used to consider how role ambiguities could affect reach, adoption, implementation, and maintenance outcomes related to rGS delivery across settings.

## Results

Participants described rGS implementation as requiring coordination across multiple clinical roles, services, and settings. Role-related tensions included but extended beyond individual tasks, reflecting broader challenges involving the fit between rGS and existing clinical routines, the distribution of responsibilities among recipients, contextual differences between Level III and Level IV NICUs, communication, and coordination of decision-making, documentation, and disclosure. Here, we describe 1) participant characteristics, 2) local care delivery adaptations; 3) workflow diagrams, and 4) role-related tensions by phase of the rGS workflow.

### 1) Participant characteristics

The study enrolled 11 participants in job task journals, and 26 participants in group workflow elicitation interviews. Twenty workflow interviews were conducted encompassing five different roles for both the Level III and Level IV systems in two rounds (10 interviews per round). Table 1 characterizes participants in both data collection strategies.

**Table 1.** Participant characteristics for workflow interviews and job task journals.

| Data Collection Strategy | NICU Level | Role | n | Years in Current Role for Each Participant | n with Any Self-Reported Genetics Training (%) |
| --- | --- | --- | --- | --- | --- |
| Workflow Interviews (n=26 participants) | IV | Nurse | 2 | 1, 15 | 0 (0) |
|  |  | Genetic counselor | 2 | 1, 2 | 2 (100) |
|  |  | Medical geneticist | 2 | 2, 18 | 2 (100) |
|  |  | Neonatologist/APP | 3 | 11, 13, 15 | 2 (67) |
|  |  | Non-genetics subspecialist | 2 | 5, 10 | 1 (50) |
|  | III | Nurse | 3 | 8, 10, 13 | 1 (67) |
|  |  | Genetic counselor | 2 | 2, 5 | 2 (100) |
|  |  | Medical geneticist | 2 | 1, 4 | 2 (100) |
|  |  | Neonatologist/APP | 6 | 3, 11, 15, 22, 24, 39 | 5 (83) |
|  |  | Non-genetics subspecialist | 2 | 2, 2 | 2 (100) |
| Job Task Journals (n=11 participants) | IV | Neurologist | 1 | 2 | 1 (100) |
|  |  | Medical geneticist | 1 | 4 | 1 (100) |
|  |  | Molecular geneticist | 2 | 2, 22 | 2 (100) |
|  |  | Genetic counselor | 1 | 2 | 1 (100) |
|  |  | Neonatologist | 1 | 4 | 1 (100) |
|  | III | Genetic counselor | 1 | 2 | 1 (100) |
|  |  | Neonatologist | 2 | 2, 31 | 2 (100) |
|  |  | Medical geneticist | 2 | 4, 12 | 2 (100) |

### 2) Local care delivery adaptations

We characterized the rGS program as relying on multiple care delivery model adaptations intended to support scale-up across settings with different levels of genetics access. These adaptations were implementation-relevant, as they redistributed rGS tasks across clinical roles and created new coordination needs for consultation, communication, documentation, and disclosure. Adaptations included: i) a consultation-based genetics service with medical geneticists and genetic counselors; ii) non-genetics subspecialists empowered to order rGS and interpret results for some patients; iii) a multi-disciplinary care team approach used as needed for complex patients; and iv) telehealth consultations to make dedicated medical genetics, genetic counseling and non-genetics subspecialist expertise available to Level III settings as needed.

Role-related challenges were then categorized according to the care delivery adaptation most directly affected by the challenge, while recognizing that many challenges were closely tied to i-PARIHS constructs, especially recipients, context, and facilitation. The categories and challenges identified were:

#### Consultation-based

Challenges related to requests for input or consultations, broadly defined, from *either* the local genetics service and/or non-genetics subspecialists (e.g. uncertainty about which service to consult, informal requests for clinical advice, questions about the ability of consulted subspecialists to adequately generate a phenotype, differences of opinion around test selection).

#### Multi-Disciplinary Teams

Challenges related primarily to the logistics of full care team dialog and decision-making (e.g. ability to reach consensus about clinical interpretation, practical aspects of team-based disclosure of results).

#### Telehealth

Challenges related primarily to remote logistical coordination of rGS processes via the telehealth service (e.g. ability to correctly route phone calls in remote facilities).

#### Other

Challenges unrelated to specific rGS program adaptations.

### 3) Workflow diagrams

The study’s final diagrams distinguish roles for reference and hospital laboratories, families, bedside nurses, genetic counselors, genetic counseling assistants, neonatologists/APPs, medical geneticists and non-genetics subspecialists. A separate diagram was also created for cases of rapid patient deterioration, added at participants’ request. From an implementation perspective, the Level III and Level IV diagrams illustrate how the same innovation (rGS) required different coordination structures depending on local context, including subspecialty availability, telehealth reliance, and mechanisms for routing results and follow-up communication. See Appendix A.

### 4) Role-related tensions by phase of the rGS workflow

Across the five workflow phases, role-related tensions in rGS centered on who had authority and responsibility to initiate, interpret, document, and disclose rGS information. These tensions were especially visible in tasks requiring close interaction between roles, and in transitions across major rGS phases. Overall, care model adaptations used to implement clinical rGS expanded the care team involved in genetic testing, redistributing work without always creating shared expectations for ownership of responsibility or communication pathways. Interview excerpts are provided in Appendix B, referenced below by excerpt number and the category of implementation challenge they were mapped to.

#### i) Identification of a patient with a suspected genetic disorder

Nursing: Bedside nurses were identified as the role with the greatest direct contact and familiarity with the patient, and the role to potentially initiate a discussion with the NICU team if something seemed unusual about a patient. Nurses indicated that raising observations about a patient required confidence that they would be listened to and not dismissed by other members of the clinical team (Excerpt 1; Other).

Medical Geneticists: While medical genetics felt empowered to manage “curbside” communication with neonatologists (Excerpt 2; Consultation), the mechanisms and responsibilities for arranging these conversations was unclear for some interviewees if a relationship didn’t already exist between clinicians.

#### ii) Initial subspecialist consultations

Non-Genetics Subspecialists: Multiple non-genetics subspecialists indicated their belief that a consultation to genetics was not always necessary if a patient’s presentation was sufficiently familiar and pertained to their subspecialty (i.e. phenotype was “single system”; Excerpt 3; Consultation).

##### Medical Geneticists

Medical geneticists recognized the logic of the single system criterion for genetics consultation (Excerpt 4; Consultation). At the same time, they noted the possibility for a non-genetics subspecialist to overlook symptoms not directly related to their subspecialty (Excerpt 5; Consultation). Some also noted that neonate phenotypes change rapidly and, as one genetic counselor put it, a phenotype is “pure [single system] until it isn’t”. Finally, medical geneticists noted the possibility of only being consulted after results were returned to clinicians to handle unexpected findings, placing a burden on geneticists to deal with an unfamiliar patient after significant time had elapsed in the diagnostic journey (Excerpt 6; Consultation).

##### Genetic Counselors

Genetic counselors expressed the desire to always be involved in both genetics and non-genetics consultation processes, noting that failure to keep them updated throughout the rGS process could result in missed opportunities, e.g. for genetic diagnosis in the event of a patient’s death (Excerpt 7; Consultation).

##### Across Roles

Several clinicians were explicit about how consultations generally tended to blur who “owned” a patient in relation to subsequent clinical tasks (Excerpt 8; Consultation).

#### iii) Test selection and ordering

Neonatologists/APPs and Non-Genetics Subspecialists: NICU providers described rGS as their default genetic test or the test best supported by existing clinical evidence for most phenotypes (Excerpts 9, 10; Consultation). Subspecialists asserted that panel tests still had value in cases where variants could be missed by rGS for context-specific reasons (Excerpts 11, 12; Consultation).

##### Medical Genetics and Genetic Counselors

Members of the genetics team described trying to achieve consensus and spell out different options for testing strategies not limited to rGS through dialog with NICU providers, without desiring to play the role of de facto “gatekeepers” or adopt a “paternalistic” attitude toward rGS ordering (Excerpt 13; Consultation).

##### Medical Genetics

Some geneticists expressed frustration with perceived pressure to order rGS (Excerpt 14; Consultation). Crucially, any genetic test ordered by the NICU (whether following the genetics team’s recommendations or not) was likely to demand “downstream” input from the genetics team to interpret results after receiving the laboratory report. A more general point of concern for medical geneticists was that the geneticist be considered a member of the care team, and their consultations recognized as providing a full genetics evaluation of a patient, not just ratification of another provider’s preferred test (Excerpt 15; Consultation).

##### Across Roles

Interviewees in diverse roles perceived that rGS was increasingly the default genetic test preferred by other NICU providers. In cases of a difference of opinion between the genetics team and non-genetics clinicians, the role of the neonatologist or APP as the responsible clinician could be in tension with the domain expertise of medical genetics. Direct conflict over test selection was described as rare but problematic when it occurred.

#### iv) Return of results from reference laboratory and clinical interpretation

##### Genetic Counselors

Particularly in telehealth, routing the laboratory results via email could be a challenge for genetic counselors because of the diversity of different clinical roles that could be involved in a case, as well as the movement of clinicians off and onto service (Excerpt 16; Telehealth). Routing result phone calls for labs sent on telehealth consultation cases was also a challenge (Excerpt 17; Telehealth).

##### Subspecialists and Medical Geneticists

In cases where rGS with unexpected complexity that were ordered without an initial medical geneticist consultation, new consultations could be initiated at this point. Medical geneticists or non-genetics subspecialists could recommend orders for additional testing at this point in the workflow (Excerpt 18; Consultation).

##### Across Roles

Clinicians lacked clarity on whose responsibility it was to document the clinical interpretation of findings at this stage (Excerpt 19; Consultation).

#### v) Disclosure of results to family and arrangement for follow-up care

##### Neonatologists/APPs

NICU providers expressed a strong desire to reserve the right to disclose themselves based on familiarity with the patient (Excerpt 20; Consultation) or to assign responsibility for disclosure to members of the care team (Excerpt 21; Consultation). Some emphasized their need to be involved in disclosure particularly for inpatient cases, while noting ambiguity of responsibility for discharged patients (Excerpt 22; Consultation).

##### Genetics and Non-Genetics Subspecialists

Geneticists and non-genetics subspecialists also justified disclosing to families based on their knowledge of the genetics and clinical implications, respectively (Excerpts 23-25; Consultation). In cases where the care team included both non-genetics subspecialists and medical geneticists, some interviewees described complex coordination and occasional miscommunication around disclosure (Excerpt 26; Consultation).

##### Genetic Counselors

Disclosures that went poorly due to miscommunication between clinicians could result in genetic counselors needing to clarify clinical information for families and provide emotional support (Excerpt 27; Consultation).

##### Across Roles

Multidisciplinary team disclosure was viewed initially by the project as an ideal to work toward. However, many clinicians expressed skepticism about the feasibility of numerous clinicians being simultaneously involved in disclosure for every case (Excerpt 28; Multi-Disciplinary Team), and concerns about the appropriateness of having clinical teams present for some sensitive conversations (Excerpt 29; Multi-Disciplinary Team).

## Discussion

This study provides insights into the complexities of clinician roles throughout a clinical rGS pathway in Level III and Level IV NICUs in which care delivery adaptations have been used to ensure a genetic test’s scalability. Our interviewees also emphasized the importance of clearly defined and expedient consultations and other processes to enable the time-sensitive decision-making often required in the NICU. From an implementation perspective, these findings suggest that scaling rGS into regular clinical practice depends not only on test availability, diagnostic utility, or reimbursement, but also on whether health systems can define and support the clinical roles, responsibilities and communication pathways needed for reliable delivery across settings.

Interviewees focused heavily on complexity arising from the simultaneous availability of consultations to both a genetics service and non-genetics subspecialists empowered to recommend genetic tests. Given that both forms of subspecialists are likely to be important in scaling genetic testing^8,16^, genomic medicine implementation may need to address the potential overlap in responsibilities between these roles. We emphasize that this overlap is not strictly a local preference—it is an implementation issue involving the fit between rGS and existing clinical routines. Of note, Mackley et al. describe an expert consensus-derived conceptual framework for understanding genetic testing workflows that have been “mainstreamed”, defined as “[shifting] all or part of the clinical genetic testing process to clinicians outside of the genetics service to facilitate patient access to genetic testing”.^28^

We found that considerations driving consultation and test selection decisions offered by our interviewees closely resembled those provided by Mackley et al. However, our empirical post-consultation workflows contrasted with the authors’ models in two key ways. First, rather than genetics and non-genetics subspecialists operating fully independently from each other, testing pathways could involve interaction between them. For example, both medical geneticists and non-genetic subspecialists could be consulted simultaneously by the NICU attending for complex cases, and support from medical geneticists could be requested by the non-genetics subspecialist if the exam or test results revealed complexity outside of their expertise.

Second, genetics and non-genetics subspecialists framed their authority over the testing process differently. Non-genetics subspecialists emphasized their own autonomy in test selection and competency in result interpretation. On the other hand, medical geneticists emphasized their roles as advisors to the NICU attending (rather than as autonomous decision-makers) while raising concerns about lack of genetics involvement in cases for which they were not consulted. The contrast between these framings of the subspecialist role highlights the importance of considerations of professional legitimacy and boundary maintenance in genetic test pathway implementation.^29–32^ Implementation strategies designed to broaden access to rGS may have unintended effects on how clinicians’ expertise is valued, as well as perceptions of ownership of clinical processes and accountability, unless role expectations are made explicit.^22^

Beyond these direct tensions over subspecialists’ roles, interviewees raised issues related to unclear responsibility around communicating and documenting when rGS tasks involved multiple clinical roles (e.g. documentation of clinical interpretation of results in the EHR after care team deliberations). Genetic testing expanded the patient’s care team, increasing the potential for diffusion of responsibility for test-related tasks, a well-known problem in intensive care.^33,34^ Diffusion of responsibility is likely to be a significant issue in settings with a complex mix of local and telehealth consultation options and/or the use of multi-disciplinary teams.^4,11,20^ From a RE-AIM perspective, lack of clarity regarding responsibility for identifying eligible patients may impact reach, while uncertainty about who should order tests or disclose results may affect adoption and implementation fidelity. Finally, excessive reliance on informal relationships to carry out rGS tasks may impede maintenance of the rGS program over time.

To address these issues, genomic medicine implementation efforts may benefit from considering the triggering conditions, sequencing and responsibilities (e.g. decision-making, communication and documentation) associated with consultations. This detailed analysis may complement higher-level conceptual frameworks, such as the one proposed by Mackley et al.^28^, for settings in which fluid interactions between attending and consulting clinicians are expected. Leveraging pre-existing clinician relationships and communication norms is likely to be an important aspect of implementation efforts.^13,35,36^ At the same time, agreed-upon consultation criteria, role-specific task expectations, standardized result-routing procedures, and shared documentation norms^37^ may need to be negotiated that preserve the benefits of local adaptation while reducing ambiguity at key workflow transitions.^38^

We note that many challenges identified here stem from tacit, local expectations that are not part of an organizational plan or clinicians’ role descriptions. Solutions to these issues may benefit from co-design approaches that leverage the perspectives of insiders (for their clinical expertise and knowledge of current practice) as well as outsiders (whose naivete creates an opportunity for insiders to publicly explain and negotiate local processes).^39^ Such co-design approaches can help clinical teams surface assumptions about authority, responsibility, and communication in the genetic testing pathway that otherwise remain implicit until a workflow breakdown occurs. In this way, workflow analysis serves not only as a descriptive method, but also as a facilitation tool for aligning implementation strategies with local context.^40^

While this study provides original insights into changes in clinical roles and responsibilities resulting from a clinical rGS program, it also has limitations. The work was conducted in two large Utah research healthcare systems. Experiences in states or countries with different regulatory environments, fewer resources, or other care model adaptations may differ. Due to logistical constraints, this study did not include the reference laboratory in workflow interviews, resulting in a limited understanding of that aspect of the rGS workflow. Finally, although this analysis was informed by implementation science frameworks, the findings should not be interpreted as an evaluation of i-PARIHS or RE-AIM. Rather, the frameworks informed the interpretation of why role-related tensions are likely to impact future efforts to scale and sustain rGS and other genetic testing pathways.

## Conclusion

Despite these limitations, our analysis helps to characterize the problem space faced by genomic medicine related to shifting clinical roles and responsibilities. Scaling genomic medicine will require implementers to understand how clinical systems can reliably coordinate the work of identifying candidate patients, consulting subspecialists, selecting tests, interpreting and documenting findings, disclosing results, and arranging follow-up. In this vein, our ongoing parent studies draw on the present analysis to inform panels of clinicians and family members engaged in co-designing strategies to enhance rGS in Level III and Level IV NICUs. By clarifying role-related barriers and facilitation needs, this work can support implementation strategies that improve the reach, adoption, fidelity, and maintained delivery of rGS while preserving the clinical flexibility needed for complex neonatal care.

## List of abbreviations

APP: advanced practice provider
EHR: electronic health record
i-PARIHS: the integrated Promoting Action in Research Implementation in Health Services framework
NICU: neonatal intensive care unit
RE-AIM: the Reach, Effectivenes, Adoption, Implementation and Maintenance framework
rGS: rapid genome sequencing
VUS: variant of unknown significance

## Declarations

### Ethics approval and consent to participate

The workflow analysis was deemed exempt by the University of Utah Institutional Review Board (IRB_00165986). All participants verbally consented to participation.

## Consent for publication

Not applicable.

## Availability of data and materials

A subset of qualitative data are available upon request. The full dataset is not available due to privacy concerns.

## Competing interests

The authors have no known competing interests.

## Funding

This work was supported by the National Institutes of Health via National Clinical and Translational Sciences Institute RC2TR004391(PIs Tristani-Firouzi and Estabrooks); and National Human Genome Research Institute K08HG013111 (PI Malone Jenkins). The content is solely the responsibility of the authors and does not necessarily represent the official views of the National Institutes of Health.

## Authors’ contributions

PT: Conceptualization, data curation, investigation, methodology, analysis, writing – original draft, writing – review and editing

CS: Data curation, analysis, writing – review and editing

LW: Data curation, analysis, writing – review and editing KAS: Writing – review and editing

RP: Writing – review and editing

JB: Methodology, writing – review and editing

TE: Writing – review and editing

MB: Data curation, analysis, writing – review and editing

MTF: Conceptualization, writing – review and editing

PE: Conceptualization, writing – original draft, writing – review and editing

SMJ: Conceptualization, writing – review and editing

## Acknowledgements

We are grateful to the clinicians in the NICUs

---

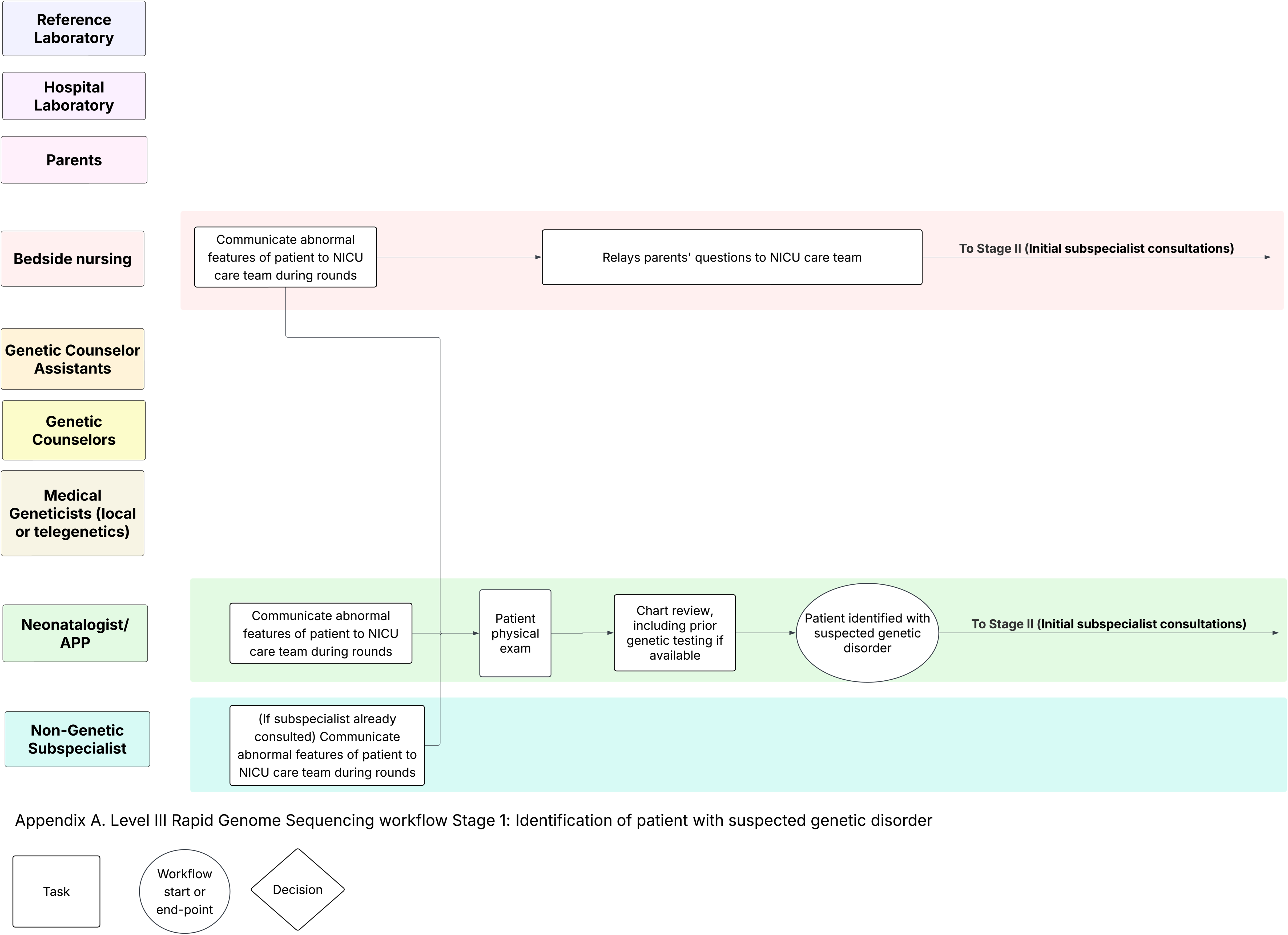

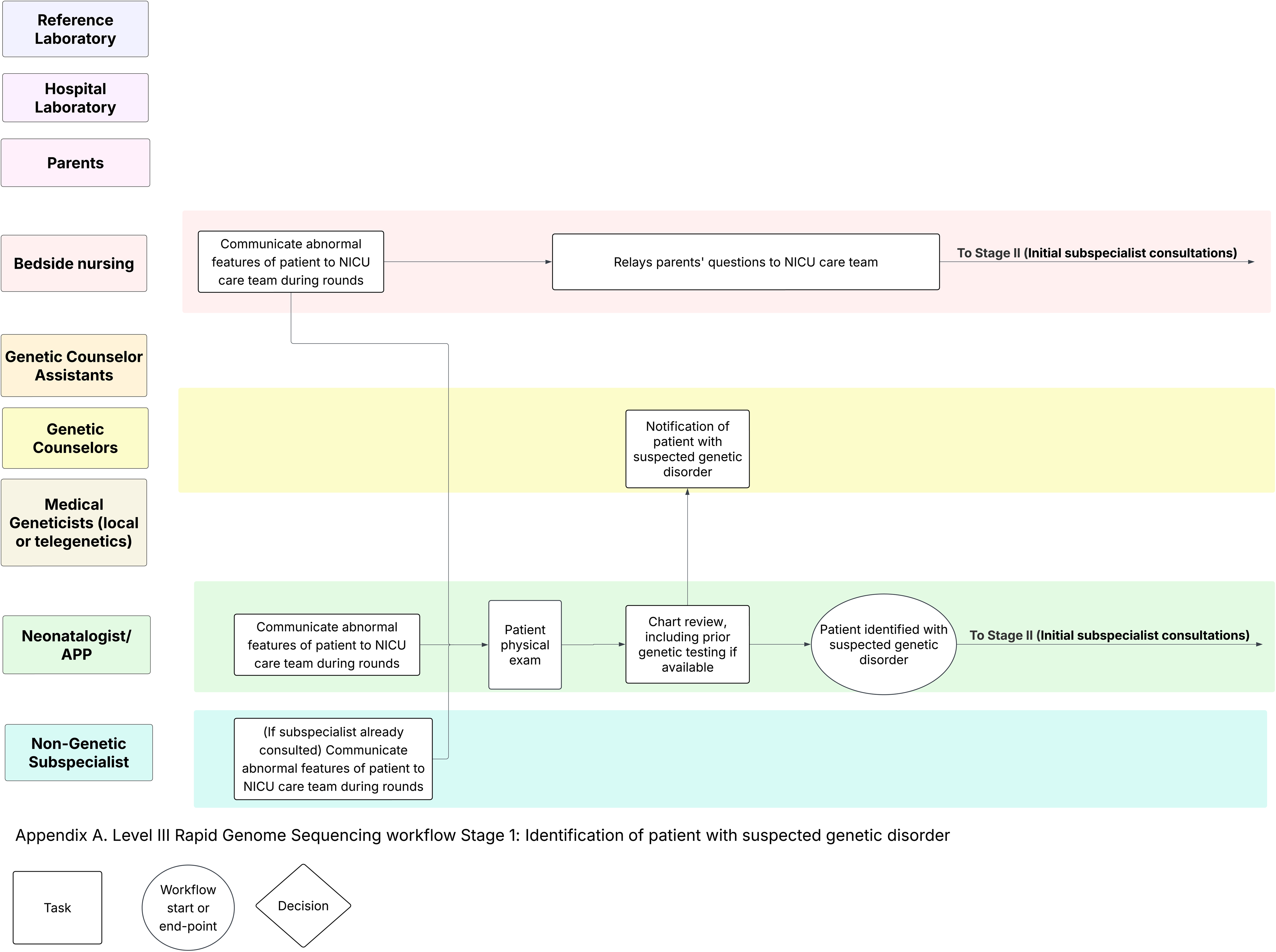

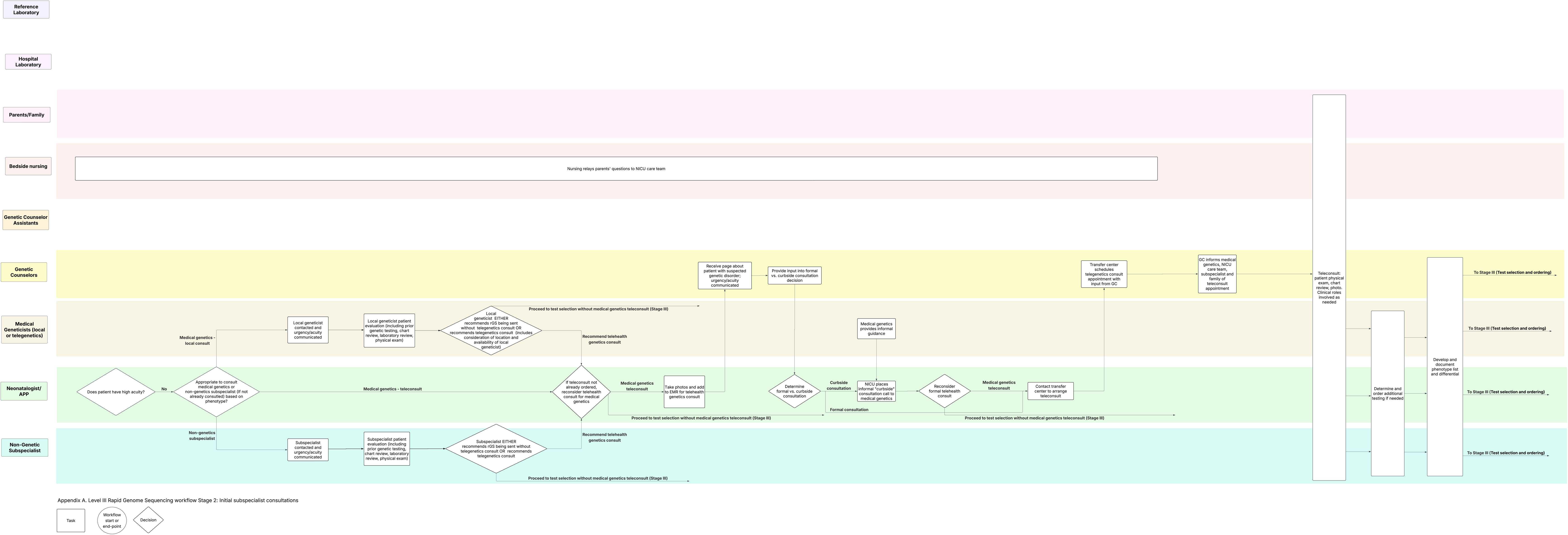

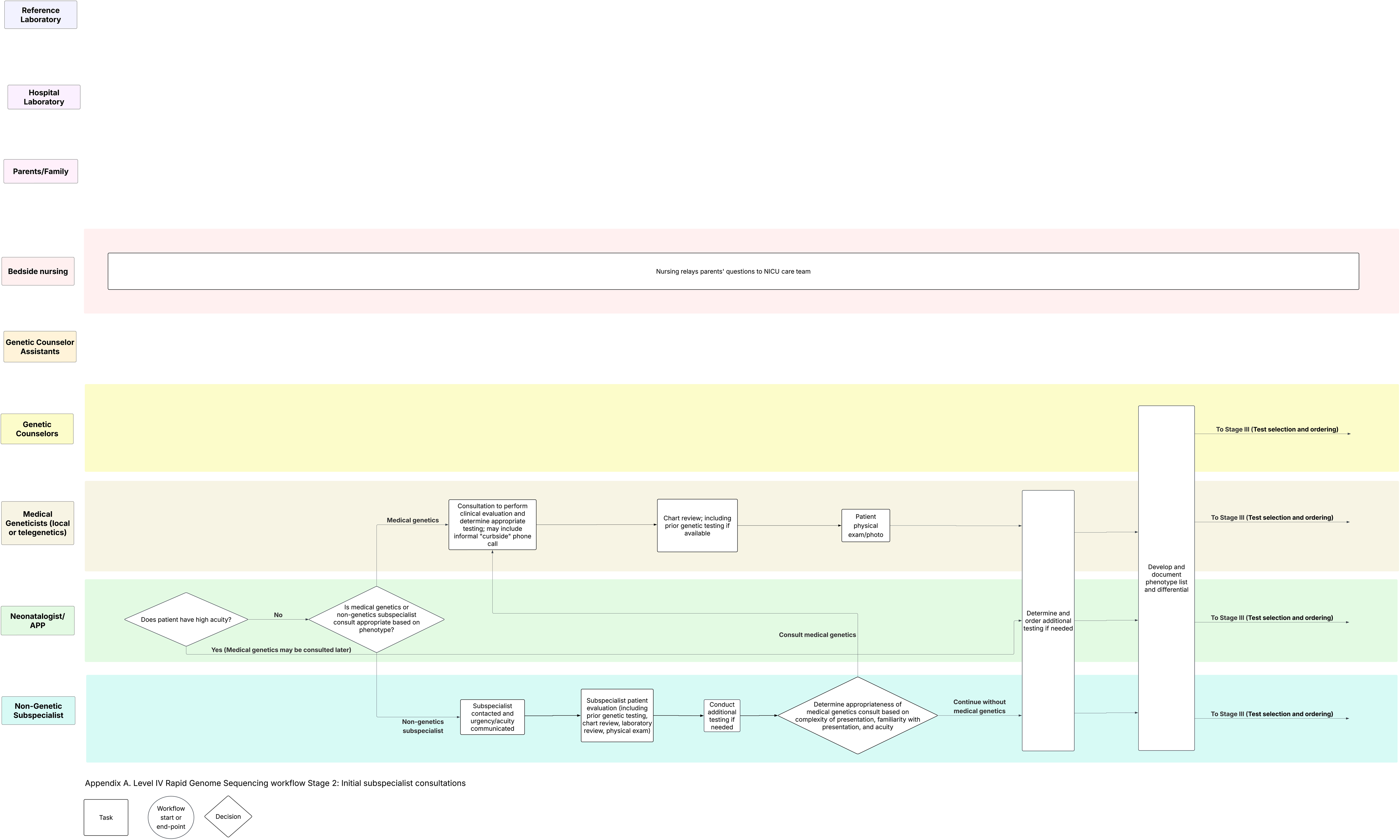

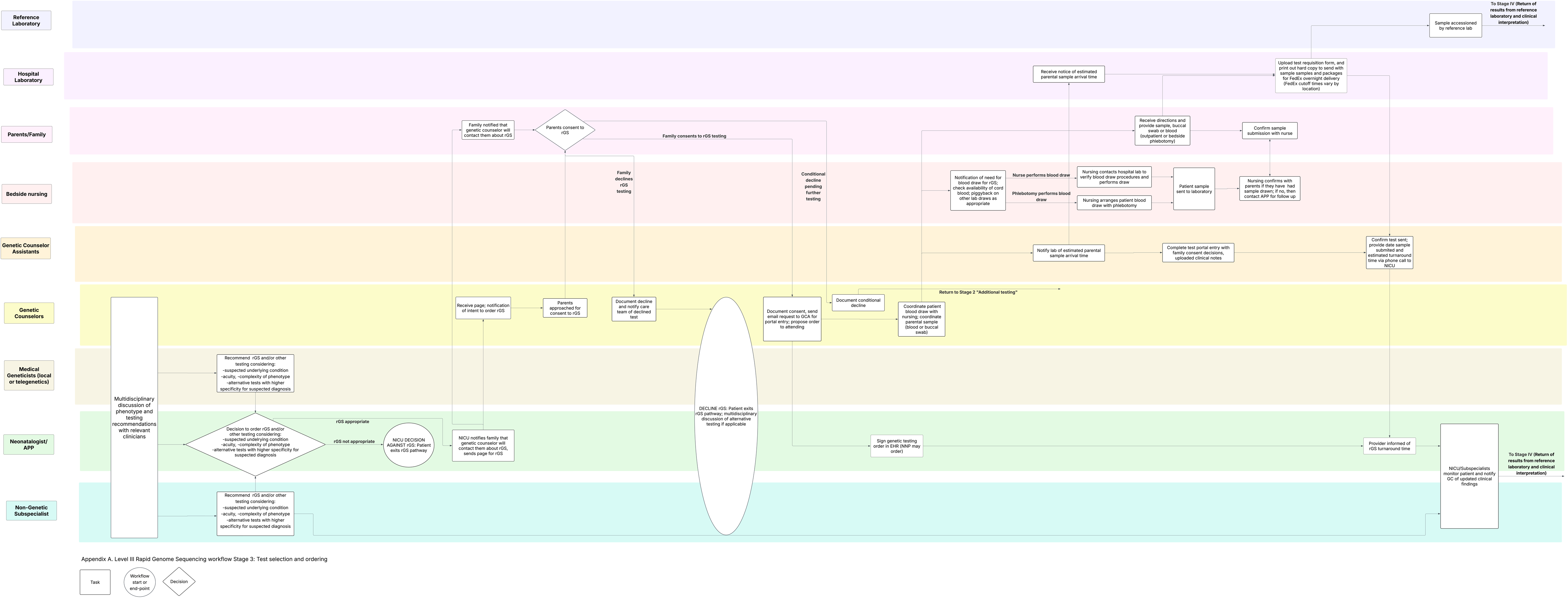

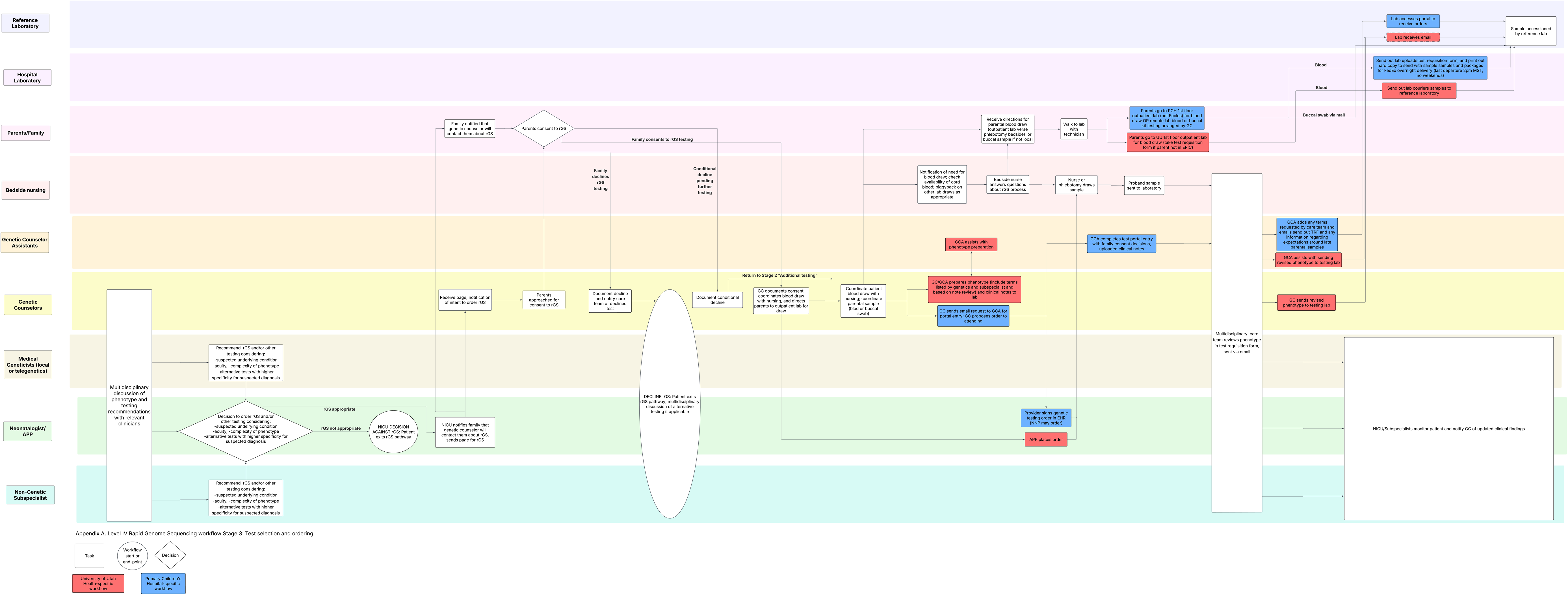

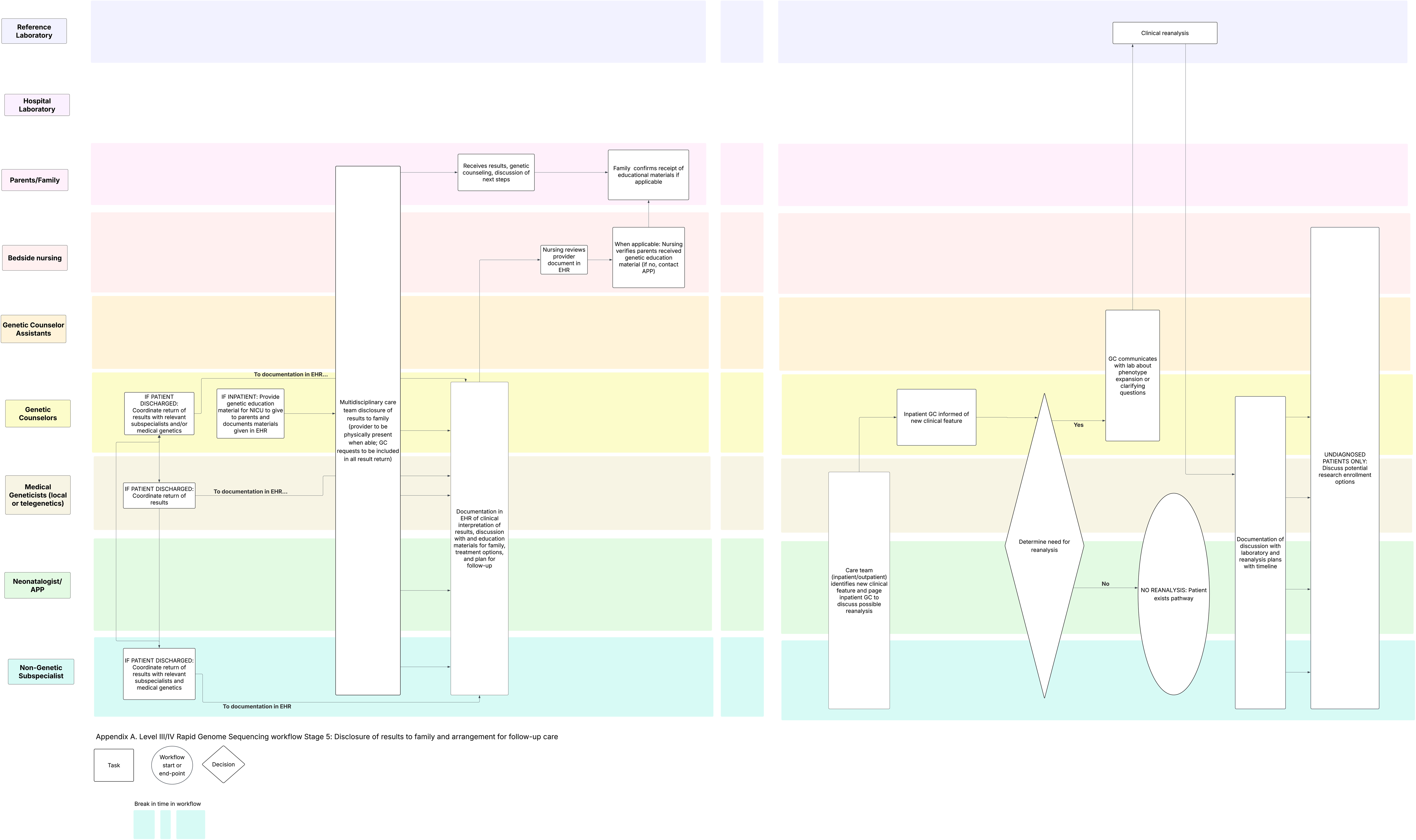

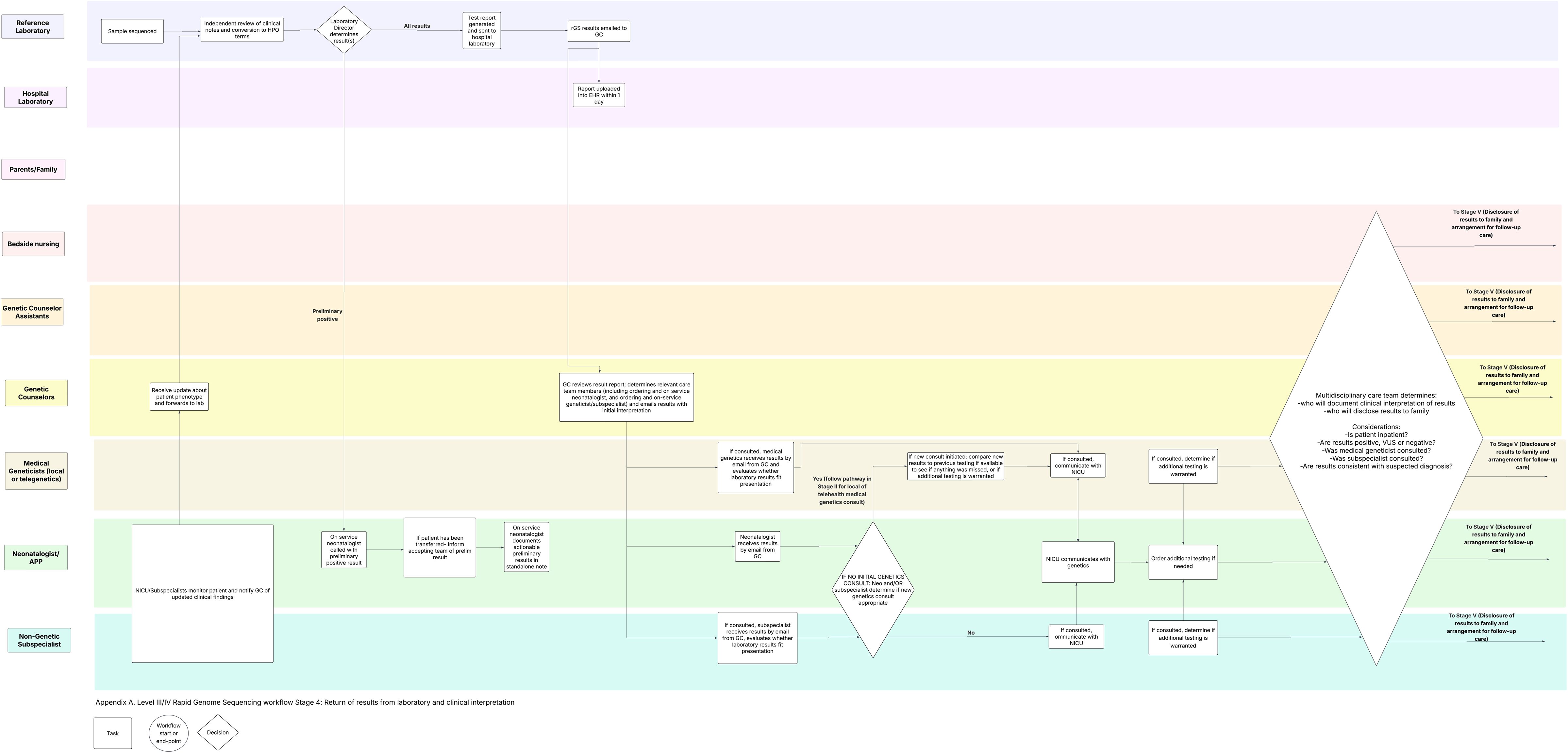

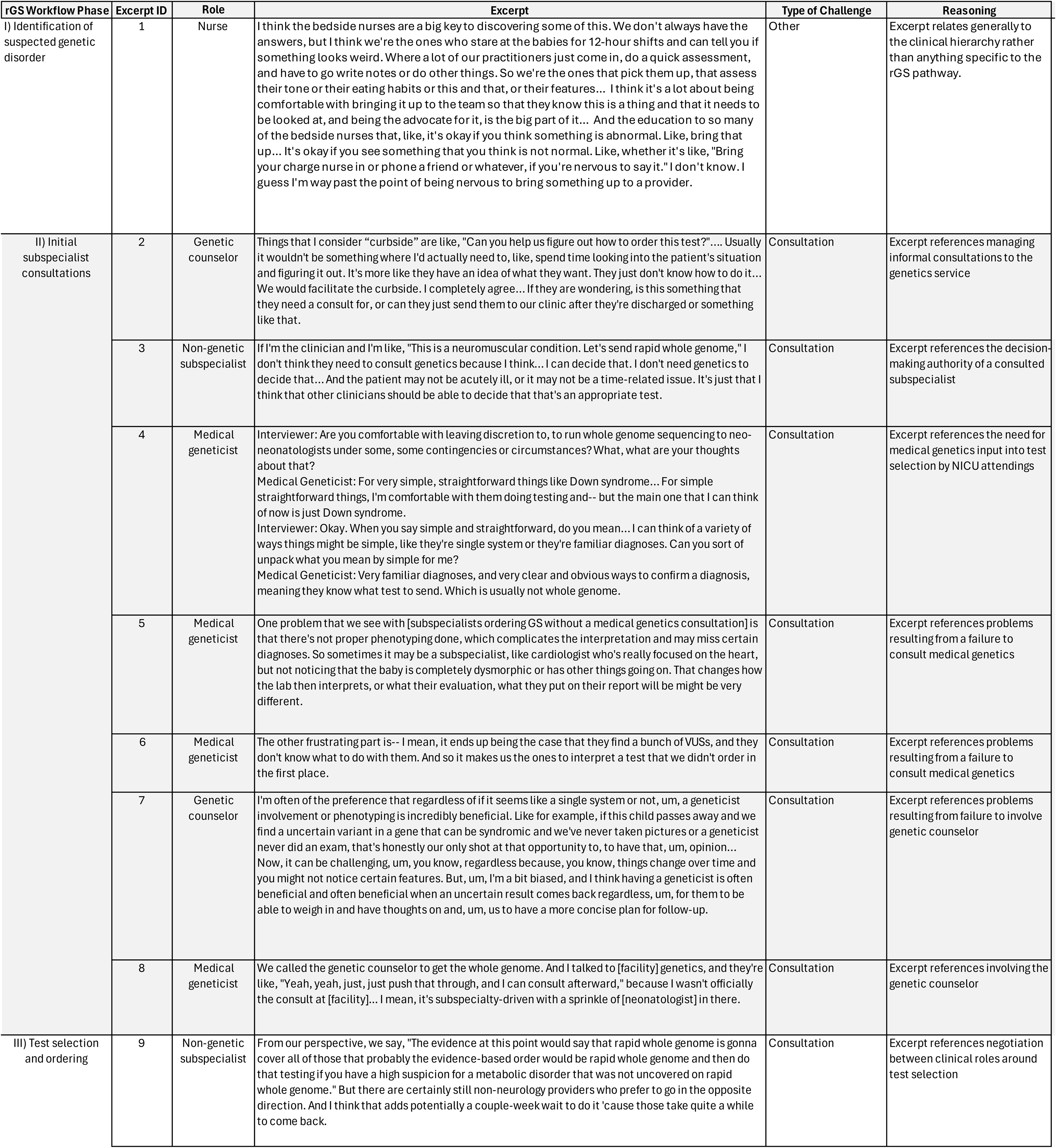
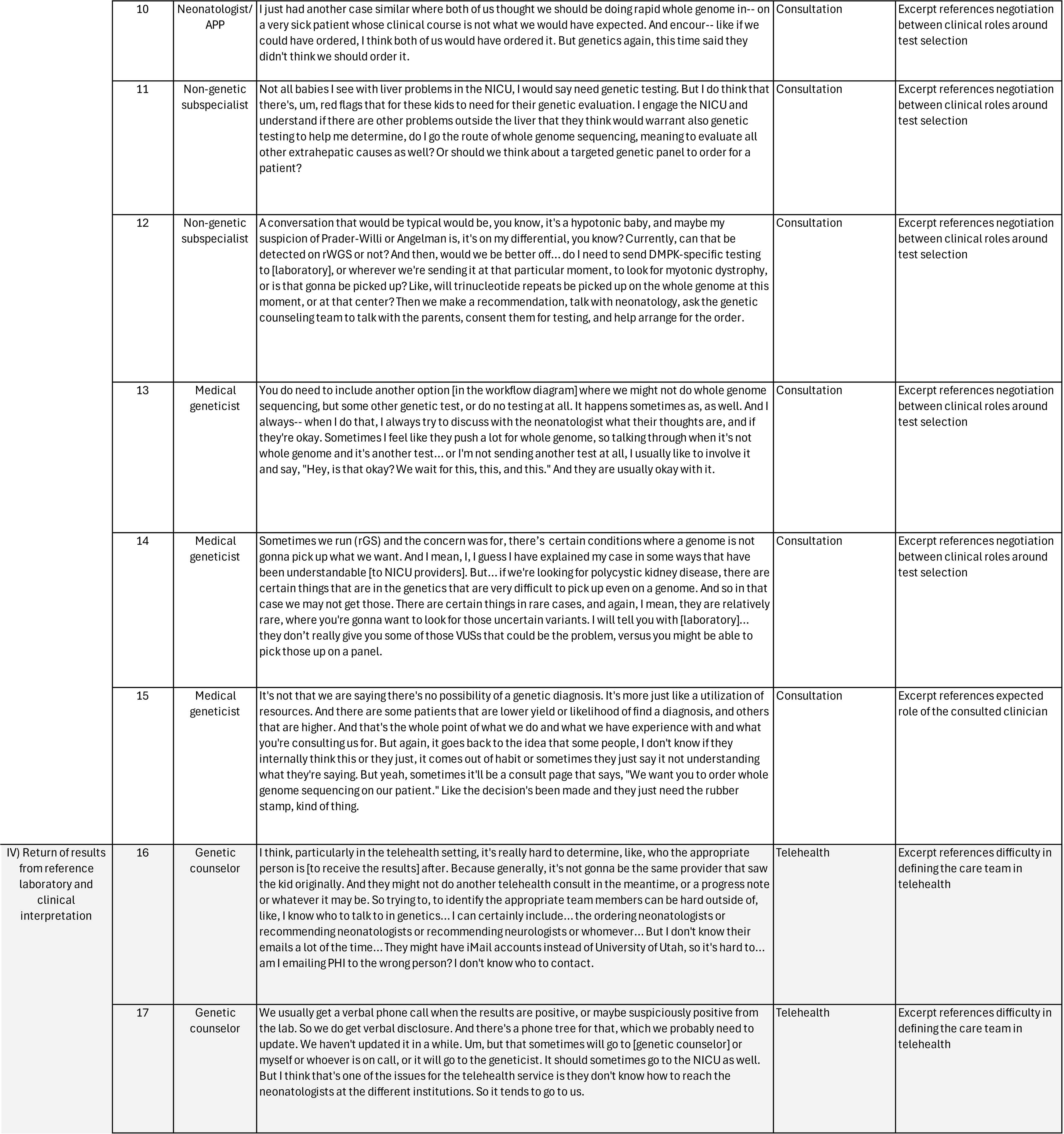
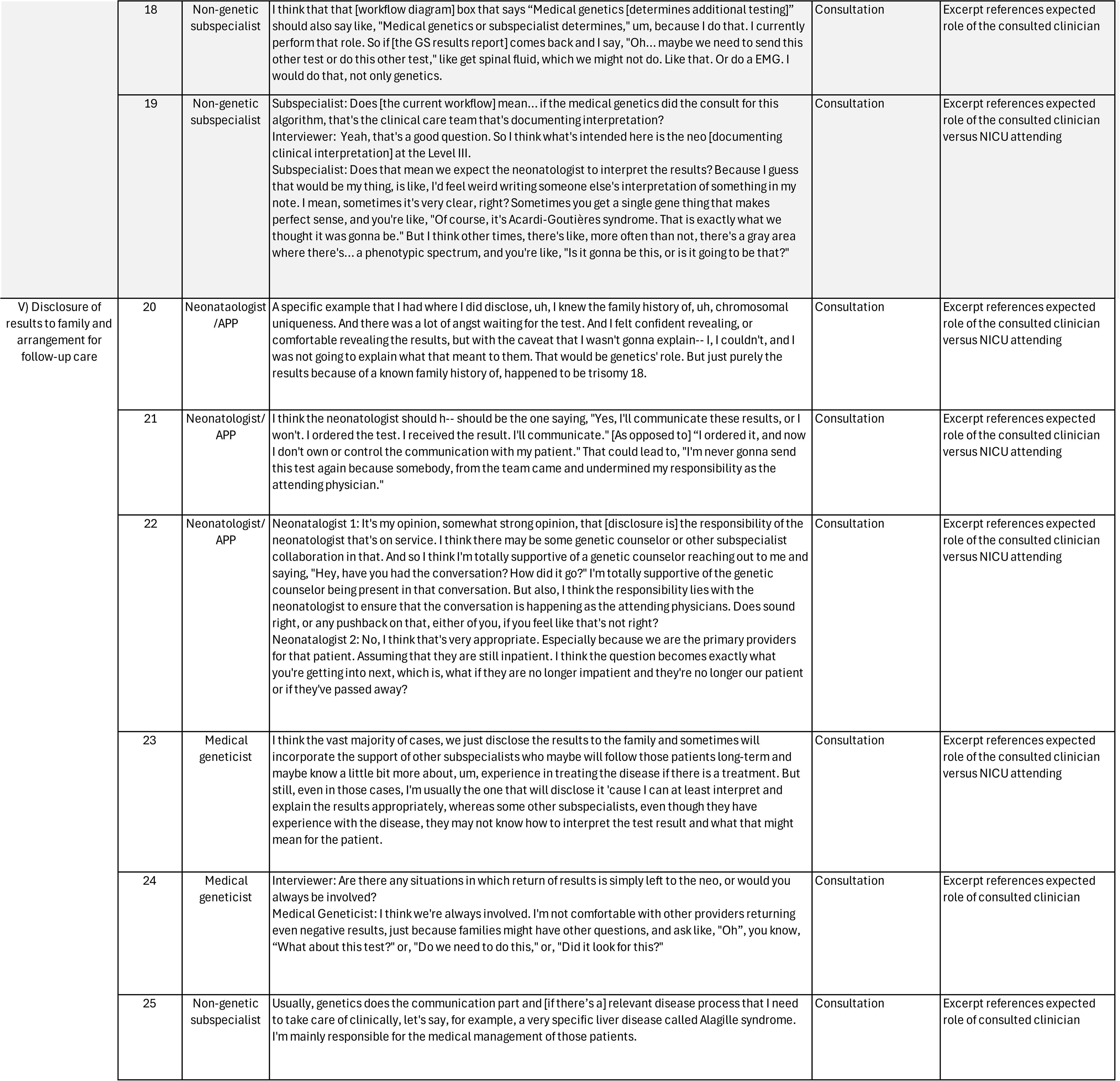
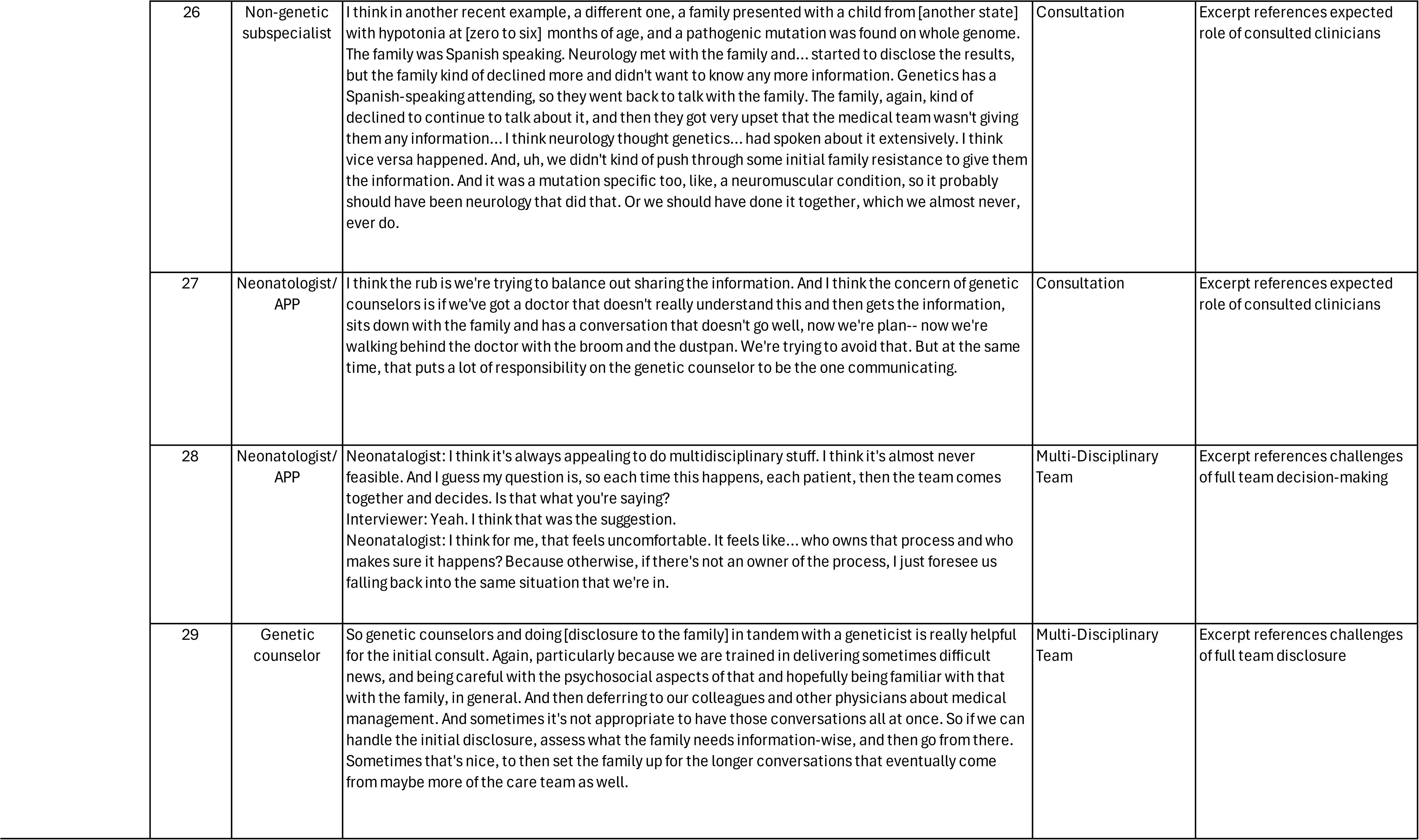

## Notes

### Competing Interest Statement

The authors have declared no competing interest.

### Author Declarations

In accordance with the Common Rule, the workflow analysis was reviewed by the University of Utah Institutional Review Board and deemed exempt (IRB_00165986).

